# The Effect of NFL and NCAA Football Games on the Spread of COVID-19 in the United States: An Empirical Analysis

**DOI:** 10.1101/2021.02.15.21251745

**Authors:** Asmae Toumi, Haoruo Zhao, Jagpreet Chhatwal, Benjamin P. Linas, Turgay Ayer

**Author notes:** **Corresponding author:** Turgay Ayer, PhD, Associate Professor, Industrial and Systems Engineering, Georgia Institute of Technology, 765 Ferst Dr., Atlanta, GA 30332.

## Abstract

**Importance:** In 2020 and early 2021, the National Football League (NFL) and National Collegiate Athletic Association (NCAA) had opted to host games in stadiums across the country. The in-person attendance of games has varied with time and from county to county. There is currently no evidence on whether limited in-person attendance of games has caused a substantial increase in coronavirus disease 2019 (COVID-19) cases.

**Objective:** To assess whether NFL and NCAA football games with limited in-person attendance have contributed to a substantial increase in COVID-19 cases in the counties they were held.

**Design:** In this time-series cross-sectional study, we matched every county hosting game(s) with in-person attendance (treated) in 2020 and 2021 with a county that has an identical game history for up to 14 days (control). We employed a standard matching method to further refine this matched set so that the treated and matched control counties have similar population size, non-pharmaceutical intervention(s) in place, and COVID-19 trends. We assessed the effect of hosting games with in-person attendance using a difference-in-difference estimator.

**Setting:** U.S. counties.

**Participants:** U.S. counties hosting NFL and/or NCAA games.

**Exposure:** Hosting NFL and/or NCAA games.

**Main outcomes and measures:** Estimating the impact of NFL and NCAA games with in-person attendance on new, reported COVID-19 cases per 100,000 residents at the county-level up to 14 days post-game.

**Results:** The matching algorithm returned 361 matching sets of counties. The effect of in-person attendance at NFL and NCAA games on community COVID-19 spread is not significant as it did not surpass 5 new daily cases of COVID-19 per 100,000 residents on average.

**Conclusions and relevance:** This time-series, cross-sectional matching study with a difference-in-differences design did not find an increase in COVID-19 cases per 100,000 residents in the counties where NFL and NCAA games were held with in-person attendance. Our study suggests that NFL and NCAA football games hosted with limited in-person attendance do not cause a significant increase in local COVID-19 cases.

**Key points:** *Question:* Did NFL and NCAA football games with limited in-person attendance cause a substantia increase in coronavirus disease 2019 (COVID-19) cases in the U.S. counties where the games were held?

*Findings:* This time-series, cross-sectional study of U.S. counties with NFL and NCAA football games used matching and difference-in-differences design to estimate the effect of games with limited in-person attendance on county-level COVID-19 spread. Our study does not find an increase in county-level COVID-19 cases per 100,000 residents due to NFL and NCAA football games held with limited in-person attendance.

*Meaning:* This study suggests that NFL and NCAA games held with limited in-person attendance do not cause an increase in COVID-19 cases in the counties they are held.

## INTRODUCTION

The Centers for Disease Control and Prevention (CDC) has advised that mass in-person events have the potential for significant spread of coronavirus disease 2019 (COVID-19)[1]. To curb the spread of COVID-19, states have implemented non-pharmaceutical interventions (NPI) with varying intensity, including closure of work places, limit on indoor and outdoor gatherings, and travel restrictions[2]. Of note, sporting events have been banned or cancelled due to the potential of mass gatherings to become “super spreader” events[3].

In early 2020, the National Basketball Association (NBA) and National Hockey League (NHL) temporarily suspended their 2019-20 seasons in an effort to limit the spread of COVID-19. A few months later, both leagues committed to resuming games in a “bubble” format where games were held in select sites with no fan attendance. The NBA suspended their season a second time due to a strike, while the NHL resumed their season with no interruption. In the late summer, the National Football League (NFL) and National Collegiate Athletic Association (NCAA) made the decision to play games in their respective stadiums, with many hosting in-person fans in a limited capacity. Despite various restrictions in place — on sporting and non-sporting events, — COVID-19 cases continued to increase nationally from 4.62 million in August 2020 to 13.84 million in December 2020; the increase in cases varied across counties and states[4].

However, the effect of limited in-person attendance in football games on the spread of COVID-19 in the hosting counties is not well-understood. Despite the recent availability of COVID-19 vaccines, strict restrictions on holding large sporting events may remain in place until a herd immunity is achieved. Hence, quantification of the effect of in-person football games attendance on the spread of COVID-19 could inform appropriate management decisions in future.

The purpose of this study was to assess whether the historical NFL and NCAA football games with limited in-person attendance have contributed to a significant increase in COVID-19 cases in the counties they were held. We used a matching method and a difference-in-differences (DID) estimator suited for time-series, cross-sectional data to estimate the effect of in-person attendance on the spread of new, reported COVID-19 cases per 100,000 residents.

## METHODS

### Data

We extracted NFL and NCAA game information, COVID-19 case counts, and COVID-19 policies and interventions from public sources. We gathered information on NFL and NCAA games played in the 2020-2021 regular season, spanning 08/29/2020 to 12/28/2020 from Pro Football Reference[5]. Extracted data included game dates, in-person attendance (yes/no), and the stadiums where the games were held. We obtained stadiums’ longitude and latitude from Wikipedia[6]. Home stadiums’ longitude and latitude were mapped to the corresponding U.S. county. We collected COVID-19 statewide non-pharmacological interventions and policies from the COVID-19 U.S. State Policy Database (CUSP) with respect to physical distance closures (closing non-essential businesses and closing restaurants except take-out), stay-at-home orders (stay-at-home/shelter-in-place and end/relax stay-at-home/shelter-in-place), and second closures and reopening (closing bars, reopening bars, reopening restaurants, and reopening non-essential businesses)[7]. We obtained county-level populations from the 2019 U.S. Census Bureau Gazetteer Files[8]. We computed county-level new cases of COVID-19 per 100,000 residents using data from the COVID-19 Data Repository by the Center for Systems Science and Engineering (CSSE) at Johns Hopkins University[9].

### Statistical analyses

We quantified the effect of interest by comparing daily changes in COVID-19 cases per 100,000 residents in counties that have held NFL/NCAA games with limited in-person attendance with those that did not hold NFL/NCAA games or have no attendance. This time series cross-sectional study used matching and a DID design to estimate the Average Treatment effect on the Treated (ATT)[10].

The overall study design consisted of three major parts: (1) constructing initial matched set for each treated county based on history of games, (2) refinement of the matched sets based on additional control variables, and (3) ATT estimation using a DID estimator.

The treatment X_*it*_ is defined as whether county *i* has had NFL and/or NCAA game(s) with in-person attendance on date *t*. If there is a game with in-person attendance in county *i* at date *t*, X_*it*_ is set to be 1. If not, X_*it*_ is set to be 0. We define post-treatment period *F* as the time period (days) following treatment, which we set to 14 days due to the incubation period of coronaviruses[11]. Similarly, we define pre-treatment period *L* to be 14 days. The outcome of interest was the change in new, reported daily COVID-19 cases per 100,000 residents from time *t* to time *t* + *F*, that is from day 0, *t*_0_, to day 14, *t*_14_.

### Constructing Initial matched set for each treated county

For each treated county X_*it*_, a set of matched counties were determined based on pre-treatment and post-treatment game history such that matched counties should have identical game history from time *t* − *L* to *t* − 1. Note that *t* is fixed for both treated and matching counties, so that with the same time trend, we can adjust for time-specific unobserved confounders later. Secondly, matched control units were excluded if they had a game with in-person attendance after time *t* but before the outcome is measured at time *t* + *F*. For each treated county *X*_*it*_ with *X*_*it*_ = 1 and *X*_*i,t*-1_ = 0, the matched set of counties is defined as:

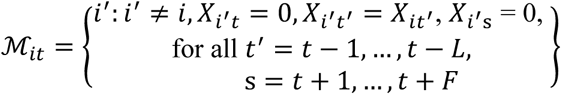

**Figure 1** provides a graphical illustration of the matching procedure. Circled zeroes stand for counties without games on that day, and circled ones mean at least one game happened. Bolded blue and red circled ones represent treatment. For illustration purposes, assume both pre-treatment *L* and post-treatment *F* are equal to 3 (days). For the bolded blue one, there is a treatment in county A on 09/04. In the first matching step, both county B and C are selected because they have the same treatment history as county A (selected with dash-dotted rectangles). Though county E also has the same history, it is not selected since it has treatment on 09/04. In the next matching step, county B is filtered out because it has a treatment within three days after 09/04. Hence, only county C is in the matched set of bolded blue treatment (with solid rectangles). In the next matching step, county B is filtered out because it has a treatment within three days after 09/04. Hence, only county C is in the matched set of bolded blue treatment (with solid rectangles). Similar matching procedures lead to no matched counties for bolded red treatment. In brief, counties are matched counties only if they first have dashed-dotted rectangles around past history and then have solid rectangles around future events.

**Figure 1:**
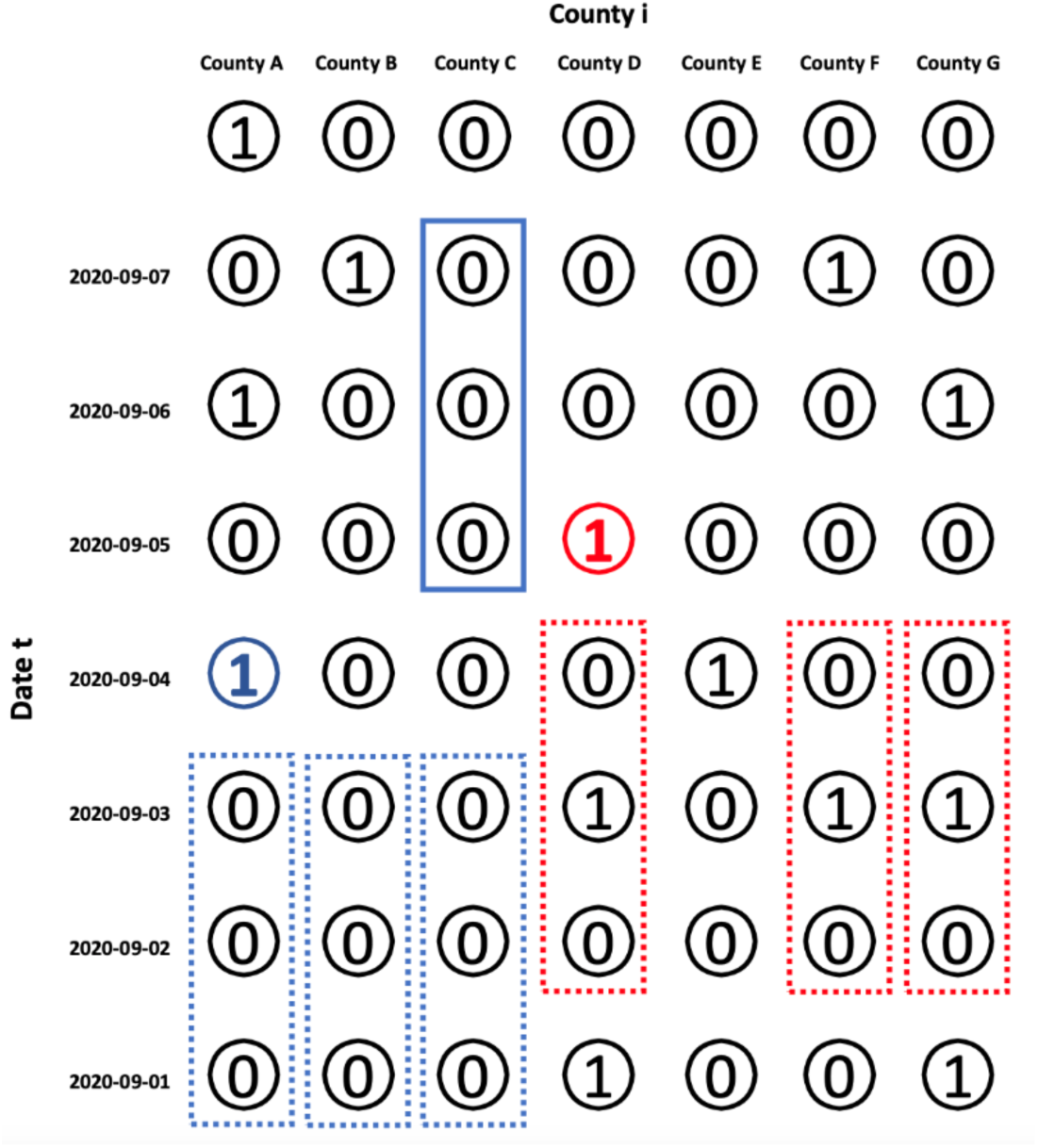
An Illustration of Matched County Set with Pre-Treatment and Post-Treatment Period Equal to Three days. *Bolded blue and red circles represent treatment observations. For each treatment, counties with the exact same pre-treatment history as the treated county are first selected (dashed rectangles). The selected counties get assigned to the matched set if their post-treatment period have no games (solid rectangles)*.

### Refinement of the matched sets based on additional controls

In the initial matching step based on history of games played, other control variables that may play an important role in spread of the disease are ignored. However, in addition to the identical history of games, we also need to ensure that treatment and control groups are similar based on factors that may influence the disease epidemiology such as population size and presence of non-pharmaceutical interventions, as otherwise the required parallel trends assumption for DID would not hold. In this step, we refine the matching algorithm to account for such control variables and ensure the validity of parallel trends assumption.

Every treated county X_*it*_ has up to 10 control counties selected from the matched set *ℳ*_*it*_ with replacement. To this end, we use the average Mahalanobis measure to calculate the distance between treated county and each matched county over time adjusting for covariates. As specified in the data section above, covariates include COVID-19 statewide non-pharmacological interventions, policies, and county-level population. Formally, the average Mahalanobis distance over pre-treatment history in our study is defined as:

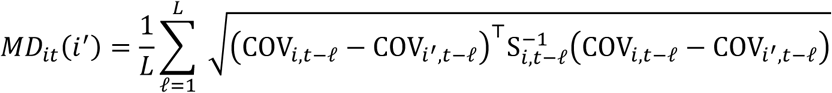

where *i*′ ∈ *ℳ*_*it*_ is a matched county of X_*it*_, COV_*it*_ is the time-varying covariates that we want to adjust for, and S_*i,t*_ is the covariance matrix of COV_*it*_. In other words, given a control county in the matched set, we compute the standardized distance using the time-varying covariates, and average it over time. For each treated county, we choose control counties with top ten smallest Mahalanobis distance, if there are any, and assign equal weights to control counties in the refined matched set. Doing so implies that up to 10 most similar control counties under Mahalanobis distance are selected for the treated county:

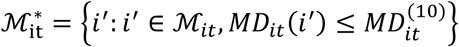

where 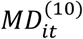 stands for the 10th smallest order statistic of *MD*_*it*_(*i*′) among the original matched set *ℳ*_*it*_.

### ATT estimation

After the refined matched sets are obtained, we calculate the counterfactual outcome using the weighted average of control counties in 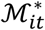. For *K* treated counties and *T* observation days, the estimated average treatment effect on the treated (ATT) of the occurrence of a football game is defined as:

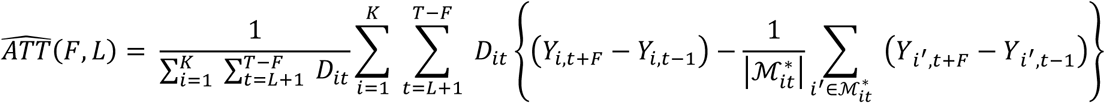

where outcome variable *Y*_*i,t*_ is the new daily cases per 100,000 residents in county *i* on day *t*, and 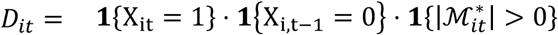 is an indicator for a treatment X_*it*_ with non-empty matching set.

Besides the time fixed effect, this model also accounts for county-level fixed effects as they are eliminated by the difference between (*Y*_*i,t*+*F*_ − *Y*_*i,t*-1_) and (*Y*_*i′,t+F*_ − *Y*_*i′,t-*1_). To compute the standard error of estimator 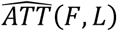 we used a block-bootstrap procedure designed for matching with TSCS data by conditioning on the weight. Number of bootstrap iterations is set to 1000.

We also performed sensitivity analyses by varying the pre-treatment period and post-treatment period. All preprocessing of data and statistical analyses were performed using R version 4.0.3 (R-Core Team, 2020). This study was deemed exempt from review by the Institutional Review Board due to the use of publicly available data.

## RESULTS

Out of 796 NFL and NCAA games played through 08/29/2020 to 12/28/2020, 528 games had in-person attendance. NCAA game attendance numbers are not available publicly. The median (IQR) attendance at NFL games through 08/29/2020 to 12/28/2020 was 11,133 (6,000 to 13,797) fans. **Figure 2** shows the distribution of NFL attendance.

**Figure 2:**
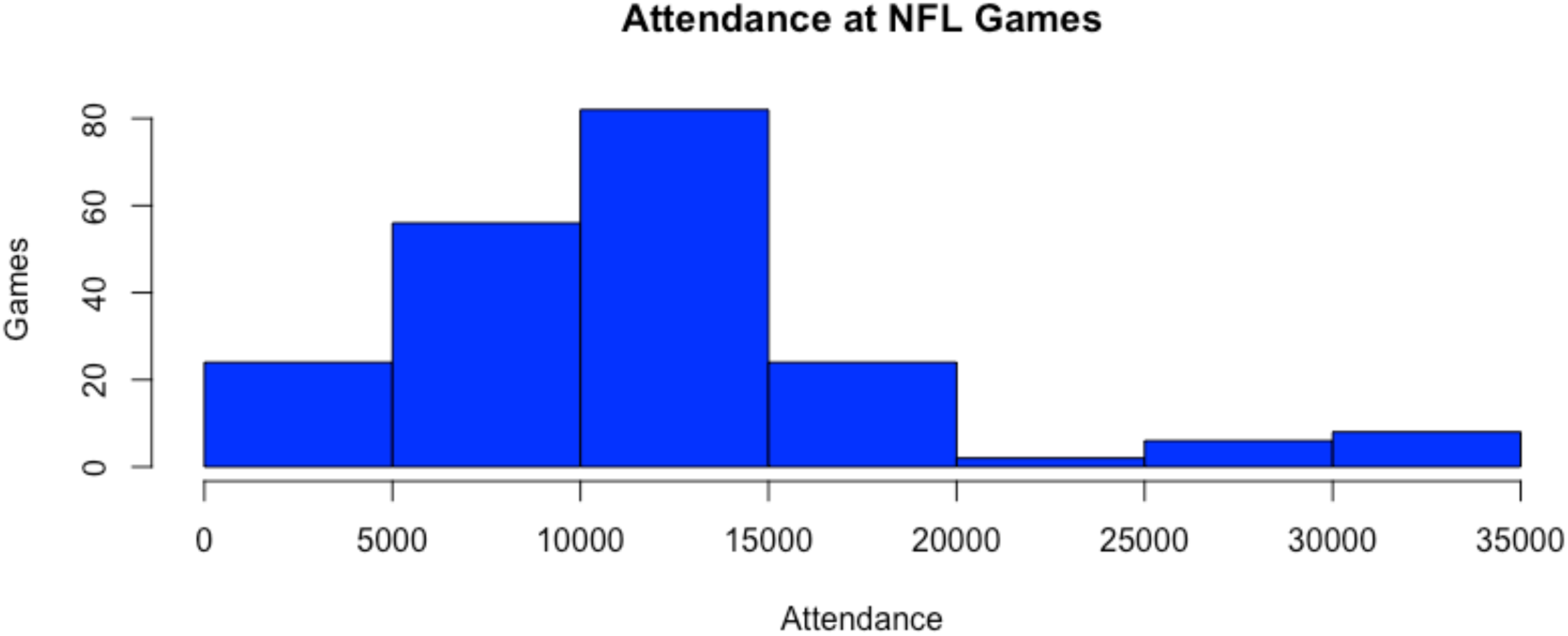
Attendance at NFL Games.

The matching algorithm returned 361 matching sets. Covariate balance in the pre-treatment time period between treated and matched control counties was assessed by computing the mean difference, measured in terms of standard deviation units, for each time-varying covariate in the pre-treatment time period. The standardized mean difference for the outcome and the other time-varying covariates stays relatively constant over the entire pre-treatment period. **Table 1** shows the estimated average treatment effect on the treated (ATT) when both post-treatment period *F* and pre-treatment period *L* are set to 14 days, along with standard error (SE) and 95% confidence interval.

**Table 1:**
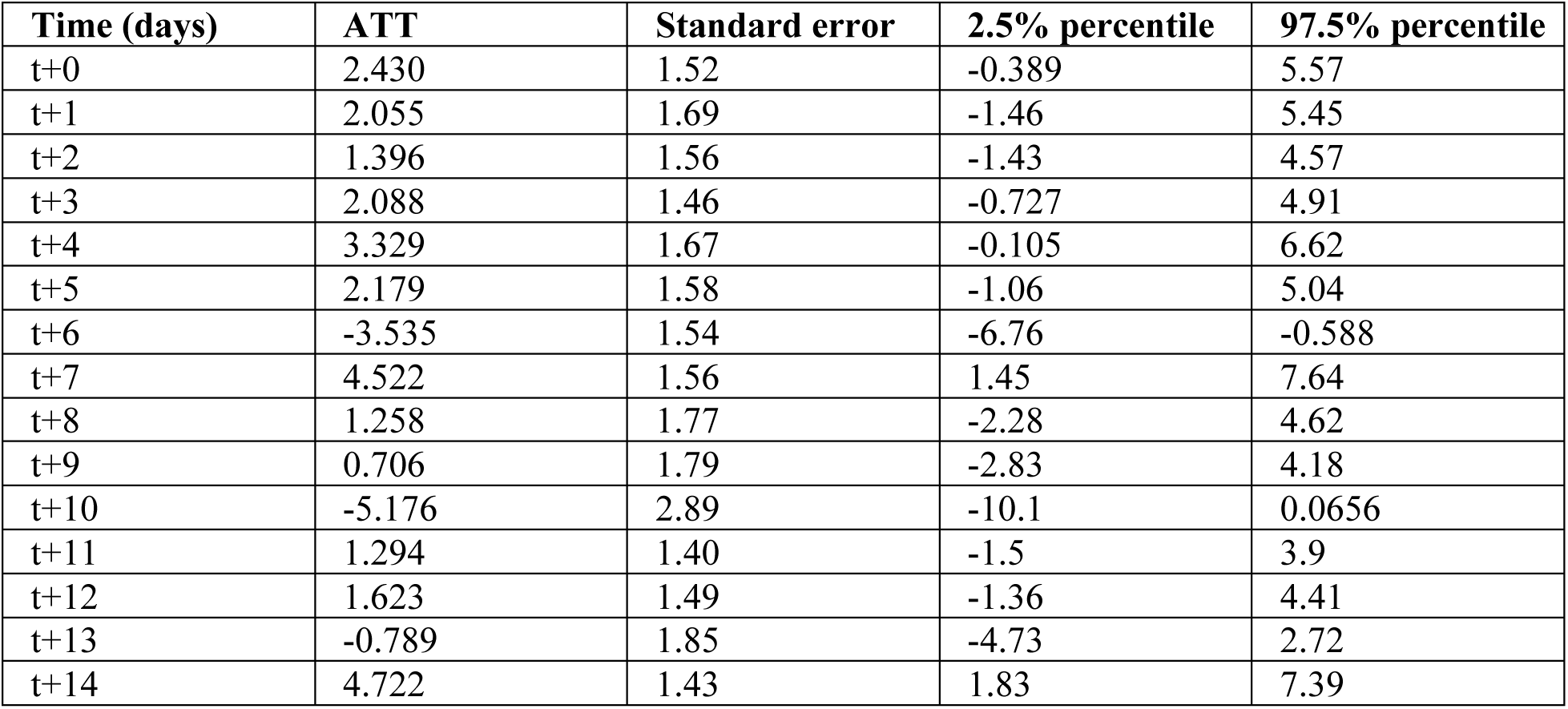
Estimated average treatment effects of NFL and NCAA football games with in-person attendance on the spread of COVID-19 over time. *The average treatment effects on the treated (ATT), standard errors (SE) and 95% confidence intervals are shown over the post-treatment period from t0 to t+14 days*.

**Figure 3** provides a graphical representation of the average treatment effects over the post-treatment period. **Table 1** and **Figure 3** suggest that the effect of in-person attendance at NFL and NCAA games is not significant, as the average treatment effects are less than 5 daily new cases of COVID-19 per 100,000 residents. At 14 days after treatment, the average treatment effect estimate in daily new cases is 4.722 per 100,000 residents.

**Table 2:**
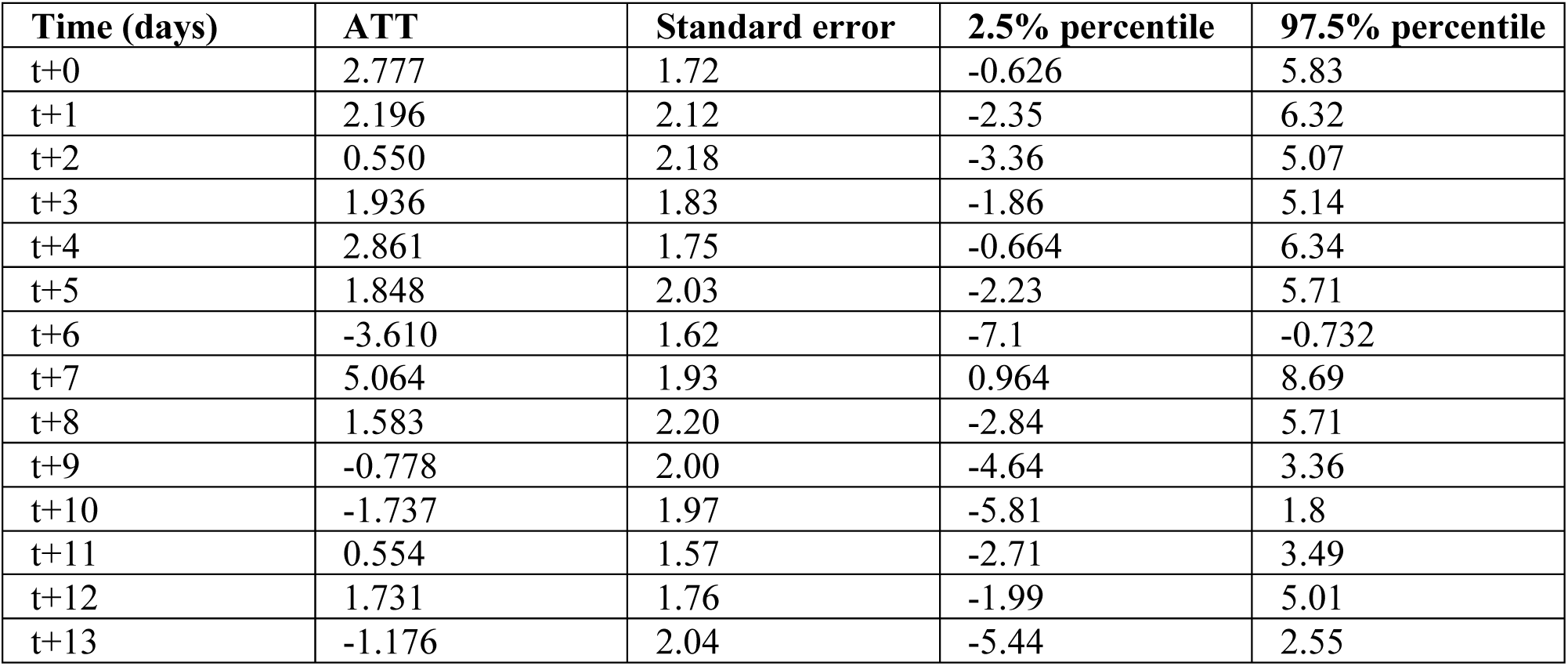
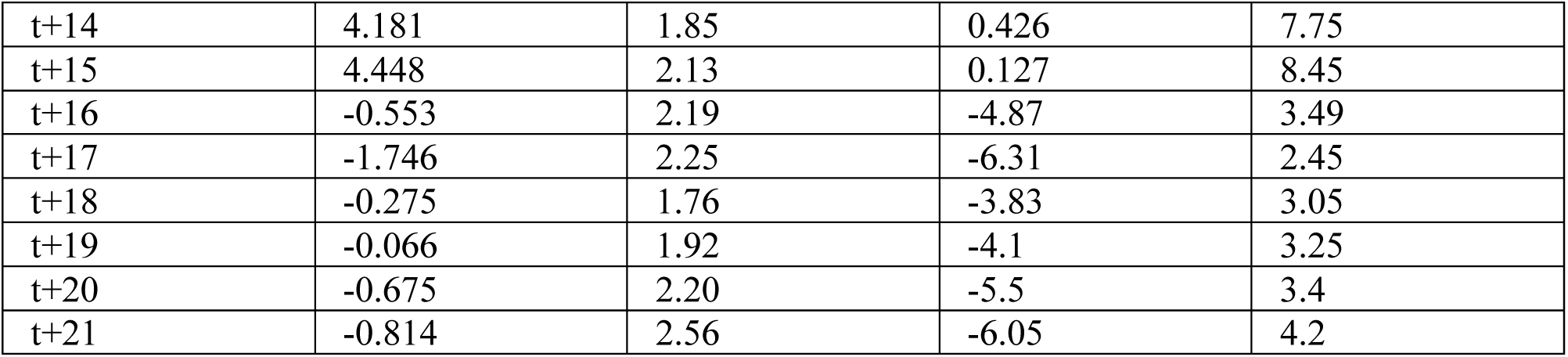
Estimated average treatment effects of NFL and NCAA football games with in-person attendance on the spread of COVID-19 over time with pre-treatment period of 14 days and post-treatment period of 21 days.

**Figure 3:**
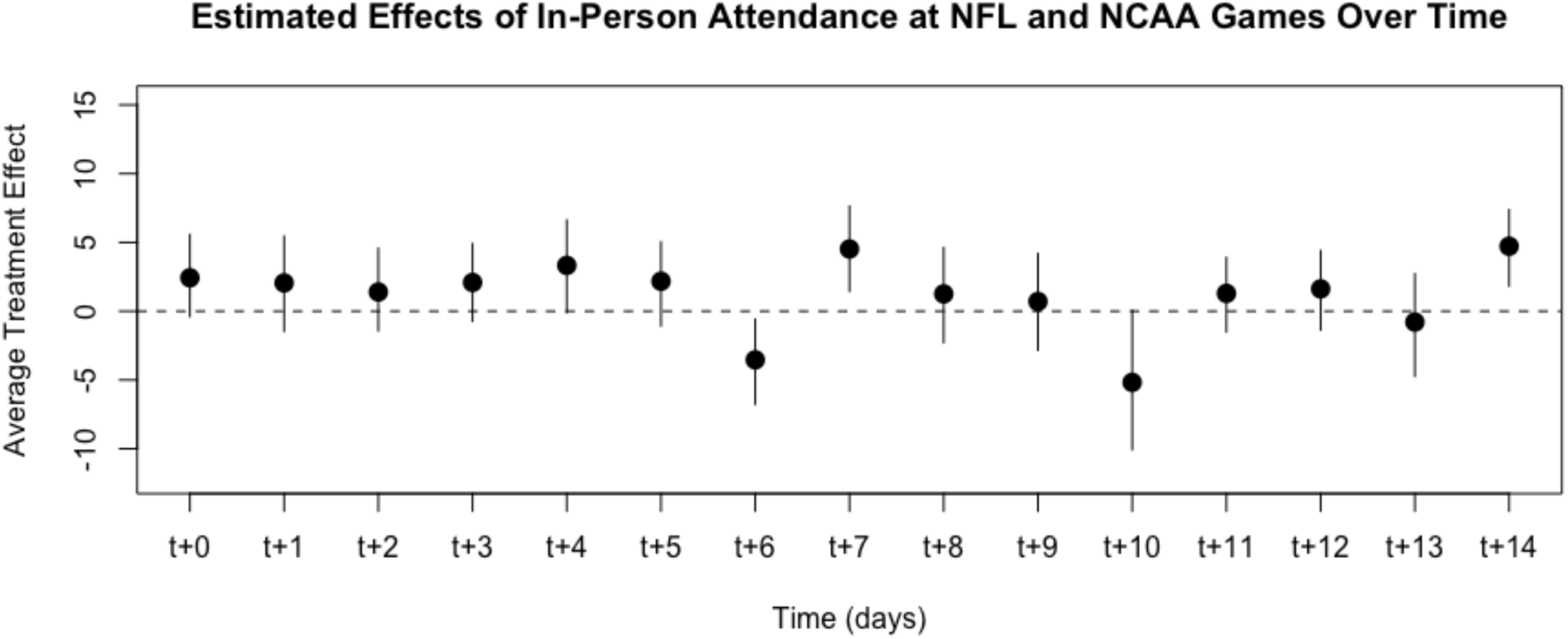
Estimated average treatment effects over time. *The average treatment effects are represented by point estimates over time (in days), with lines corresponding to standard errors*.

To evaluate the robustness of our results, we performed sensitivity analyses by varying the pre-treatment period and post-treatment period parameters (*L, F*) to (14, 7), (14, 14), (14,7) and (21,14) days. Across the different (*L, F*) parameter specifications, we found that the average treatment effects insignificant for the entire post-treatment period as they do not surpass 5 daily new cases of COVID-19 per 100,000 residents. Table 2 shows the estimated average treatment effect on the treated (ATT) with (*L, F*) set to (14, 21), along with standard error (SE) and 95% confidence interval.

## DISCUSSION

In this time-series, cross-sectional study of U.S. counties with NFL and NCAA football games, we used matching and a difference-in-differences estimator to estimate the effect of games with in-person attendance on county-level COVID-19 spread compared to games held without any fans. The study considered the effect of both NFL and NCAA games since many counties had both an NFL or NCAA game in the pre-treatment or post-treatment periods, hence the effect of NFL games alone or NCAA games alone could not be studied separately. Games with in-person attendance were matched with a set of control counties that share the same game history from time *t* − 14 to *t* − 1 days. We selected 14 days based on numerous papers that found the incubation period for COVID-19 to be 2 to 14 days[11]. To control for post-treatment bias, we apply the idea of marginal structural models by excluding control counties that had a game with in-person attendance after time t but before the 14-day post-treatment period is complete. We found that the average treatment effects on the treated of in-person attendance at NFL and NCAA games on new, daily reported COVID-19 cases (per 100,000 residents) was not significant over a 14-day period as they did not surpass 5 new cases per 100,000 residents. We surmise that the NFL and NCAA policies with regards to limited in-person attendance, mask use and social plus physical distancing measures in stadiums does not lead to a significant increase in the community spread of COVID-19[12, 13]. Additionally, an important number of NFL and NCAA football stadiums are outdoors or possess a retractable roof, which could’ve had an impact on mitigating spread[14].

There are limitations to this study. The Stable Unit Treatment Value Assignment (SUTVA) assumption stipulates that each unit receives the same form or version of the treatment[15]. The treatment in this study, namely in-person attendance, was defined as a binary (yes or no fans) and therefore the results do not capture the possible impact of differing attendance numbers (**Figure 2**) on the outcomes. When the NCAA attendance numbers are made publicly available, additional sensitivity analyses can be performed to establish whether varying attendance numbers have an impact on COVID-19 spread. We also did not control for other large gathering events, such as political rallies, as some have been found to cause a local spread in COVID-19 cases[16]. We also did not account for the spillover effects to the counties adjacent to the ones hosting NFL/NCAA games. Finally, our analysis could not assess whether the Super Bowl LV game held on February 7^th^, 2021 in Tampa, Florida is associated with an increase in the spread of COVID-19 cases because of post-game celebrations.

Our study provides new information on the effect of football games with in-person attendance on COVID-19 spread. This time-series, cross-sectional study with difference-in-differences design suggests that NFL and NCAA games held with limited in-person attendance do not cause an increase in COVID-19 cases in the counties they are held. Further research is needed to account for spillover effects to counties adjacent to the ones hosting games. While COVID-19 vaccination has started, restrictions will likely stay in place for several months and possibly until the start of the 2021-22 NFL and NCAA seasons. Our study provides evidence suggesting that in-person attendance of football games with social distancing and mask use could be resumed in the 2021-22 season.

## Data Availability

The data that support the findings of this study are available on request from the corresponding author, AT.

